# Psychological and social impact of COVID-19 in Pakistan: Need for Gender Responsive Policies

**DOI:** 10.1101/2020.10.28.20221069

**Authors:** Fauziah Rabbani, Hyder Ali Khan, Suneel Piryani, Areeba Raza Khan, Fahad Abid

## Abstract

**BACKGROUND:** COVID-19 has rapidly crossed borders, infecting people throughout the world. Women may be especially vulnerable to depression and anxiety due to the pandemic,

**AIMS:** This study attempted to assess how gender impacts risk perceptions, anxiety levels behavioral responses to the COVID 19 pandemic in Pakistan in order to recommend gender responsive health policies

**METHODS:** A cross-sectional online survey was conducted. Participants were asked to complete a sociodemographic data form, the Hospital Anxiety and Depression Scale (HADS), and questions on their risk perceptions, preventive behavior and information exposure. Regression analysis was used to assess effects of factors such as age, gender and household income on anxiety levels.

**RESULTS:** Of the 1390 respondents, 478 were women, and 913 were men. Women considered their chances of survival to be relatively lower than men (59 % women vs 73% men). They were also more anxious (62% women vs 50% men), and more likely to adopt precautionary behavior, such as avoiding going to the hospital (78% women vs. 71% men), not going to work (72% women and 57% men), and using disinfectants (93% women and 86% men). Men were more likely to trust friends, family and social media as reliable sources of COVID-19 information, while women were more likely to trust doctors.

**CONCLUSION:** Women experience a disproportion burden of the psychological and social impact of the pandemic compared to men. Involving doctors in healthcare communication targeting women, might prove effective. Social media and radio programs may be effective in disseminating information related to COVID among men.

## Background

The occurrence of novel Coronavirus Disease 2019 was first detected in December, 2019 in Wuhan, China (1). On March 11, 2020, the World Health Organization declared the outbreak of the disease as a global pandemic. Within two weeks of this announcement, COVID-19 cases outside of China had increased 13-fold with the number of affected countries rising by 3-fold (2). Since its emergence, there are more than 15.2 million confirmed global cases of the virus with the number of deaths exceeding 623,000 as of 23July, 2020. In the initial stages of the outbreak, most cases of COVID-19 that were exported internationally had a history of prior travel to Wuhan (3). Despite its close geographic proximity with China and Iran, the first two cases of COVID-19 in Pakistan were reported on February 26, 2020 (4). To curb the spread of the virus, Provincial Governments in Pakistan initiated partial and subsequent, complete lockdowns in their respective administrative territories. These measures however, were taken in phases, with educational institutions across the country closing on 13 March, 2020 in response to the pandemic (5). As of 23 July, 2020, Pakistan has over 260,000 confirmed cases of COVID-19, and approximately 5,700 deaths (1).

Exposure to a traumatic event, such as a global health crisis, is associated with an increased incidence of anxiety and depression (6). Moreover, the stigma and isolation due to infectious diseases could generate anxiety (7). A study conducted on a sample of Severe Acute Respiratory Syndrome (SARS) survivors in Hong Kong revealed increased levels of psychological distress and anxiety, not only during the epidemic but also one year following the outbreak (8). Another research concluded that SARS had long-term psychiatric effects on survivors, with post traumatic stress disorder (PTSD) and depressive disorders being the most prevalent conditions recorded (9). Moreover, a US survey conducted during the H1N1 pandemic, with 7236 participants, suggests an increase in the prevalence of anxiety (10). Similarly, one internet survey conducted in China (n=7,236) reported that over one third participants exhibit symptoms of anxiety disorders during the COVID-19 outbreak while, one fifth recently experienced sleep issues and depressive symptoms (11).

Past research during large epidemics/pandemics, such as during the Ebola outbreak of 2014-2016in West Africa suggests that women may be disproportionately impacted during public health emergencies. The COVID-19 pandemic is also expected to affect women’s health and increase their short-term and long-term needs for livelihood support, and health (12). One study in the heavily impacted areas around Wuhan, highlighted an increase of 7% in prevalence of posttraumatic stress symptoms (PTSS), and women had significantly higher levels of PTSS than men (13). One study conducted in Turkey revealed that the pandemic had a greater psychological impacted on women than men. In this study of 343 respondents, women had significantly higher depression and anxiety scores (14). During the SARS epidemic, people with higher levels of anxiety were frequently found to adopt precautionary behaviours (15). Studies conducted in Singapore and Hong Kong also report that women were more likely to adhere to precautionary behaviours during SARS than men (16). Similarly, women in the UK were found to practice precautionary behaviours such as hand washing and disinfecting surfaces more often than men (17).

Female responses to stress and trauma may be contributing factors towards anxiety. Studies also indicate that gendered responses to trauma contribute to the greater onset of depression and PTSD in women (18,19). Evidence attributes this to women’s belief that worry is uncontrollable, and may cause anxious thoughts (20). The manner in which young boys and girls are socialised into their gender roles has an impact on these perceptions. A review discussed how mothers are more likely to converse about their emotional condition with their daughters as compared to with their sons (21). Further, young boys are conditioned to exercise problem-solving skills for managing their emotions, girls are traditionally granted less autonomy. This increases their dependency on others, and reduces the capability to effectively cope with anxious thoughts (21).

This study aims to assess gender differences in perceived risk, anxiety levels and behavioural responses to COVID 19. This study uses sociocultural patterns to examine the effect of age, gender, and household income on anxiety levels, and develops gender responsive policy options for grappling with the pandemic in Pakistan.

## METHOD

### Study design

This is a cross-sectional study, with the survey tool disseminated online. The survey in Pakistan was conducted eleven weeks after the first case was reported on February 26, 2020; during a government-imposed lockdown.

### Study Participants

A convenience sampling strategy was used to recruit participants. The questionnaire was launched for two weeks on the social media pages of a Karachi based university hospital. Potential study participants were encouraged to share the link on their social media platforms. People aged 18 and above, residing in Pakistan for the last month, with access to internet and willingness to participate in the study were included. Participants who could not respond to the study tool in either English or Urdu, and those who reported having filled the questionnaire at least once before were excluded. A total of 1406 respondents filled the online questionnaire. Fifteen respondents preferred not to disclose their gender. Thus 1391 participants were included in this study.

### Data Collection

Data was collected through an online self-administered structured questionnaire developed on Google Forms. Respondents were inquired about their gender, age, level of education, household income, and city of permanent residence. They were asked how likely it is that they or their families might be infected with COVID-19 if no preventive measures were taken. Further questions assessed how participants perceived the severity of the symptoms caused by COVID-19, their likelihood of survival if infected, and their adoption of precautionary measures. Respondents also rated the reliability of various sources of COVID-19 information. Subsequently, the psychological impact of COVID-19 on respondents’ job, personal life, sleep pattern, and eating habits was assessed. Participants’ anxiety and depression levels were assessed using the validated Hospital Anxiety and Depression Scale (HADS). This consists of 14-items (7-items of anxiety and 7-item of depression) scored on a four-point Likert scale. The lowest possible scores for anxiety and depression are 0, and the highest possible score is 15 for anxiety and 21 for depression. The scale defines a normal score as≥ 7, borderline abnormal score as 8-10, and abnormal score as ≤11. Higher scores imply the severity of anxiety or depression (22– 24).

### Data Analysis

Data collected from respondents was stored in Google Spreadsheets then imported to Microsoft Excel and SPSS version 21 (IBM Corp). Data was cleaned, coded, and analysed using SPSS Version 21. A descriptive analysis was performed. Results were tabulated as number (percentage) for qualitative variables and mean (±standard deviation) for quantitative variables. Independent t-test or Mann Whitney U test or Pearson Chi-square test was applied to assess the differences between women’s and men’s perception of susceptibility and severity towards COVID-19, anxiety and depression, psychological impact of COVID-19, adoption of precautionary measures, and reliability of information sources. Bivariate and multivariate binary logistic regression analyses were performed to identify predictors (age, gender, and household income) of anxiety and depression. Initially, a single predictor at a time was entered; crude odds ratio (OR) and 95% confidence interval (CI) were computed using bivariate analysis. Multivariate analysis with all predictors entered at the same time was completed to adjust for the effect of confounding, and adjusted OR and 95% CI were computed. All statistical tests were two sided and p-value of ≤0.05 was considered statistically significant.

### Ethical Considerations

Ethical approval was obtained from the Ethical Review Committee (ERC) of the Aga Khan University, Pakistan. Prior to filling the online questionnaire, each respondent was asked to provide consent for participation in the survey.

## Results

A total of 1391 participants’ responses were included in the analysis. Table 1 shows the sociodemographic characteristics (age, education, and household income) of women and men which are comparable. Majority of the respondents were aged between 25-34 years (69% women and 66% men) and possessed a Bachelor’s Degree or above (75% women and 79% men). About 29% of women and 17% men preferred not to disclose their household income. Around one-third of women (32%) and two-fifths of men (40%) mentioned that their household income is below PKR 60,000. More women (69%) than men (40%) from Karachi participated in the survey (Table 1).

**Table 1.** Sociodemographic Characteristics of the Respondents by Gender.

| **Characteristics** | **Female**<br>**n= 478**<br>**No. (%)\*\*** | **Male**<br>**n=913**<br>**No. (%)\*\*** | **P-value‡** |
| --- | --- | --- | --- |
| **Age** |  |  |  |
| 18-24years | 167 (34.9) | 291 (31.9) | 0.833 |
| 25-34 years | 163 (34.1) | 315 (34.5) |  |
| 35-44 years | 89 (18.6) | 185 (20.3) |  |
| 45-54years | 32 (6.7) | 72 (7.9) |  |
| 55-64 years | 19 (4.0) | 32 (3.5) |  |
| 65-74 years | 7 (1.5) | 16 (1.8) |  |
| 75 years or above | 0 | 1 (0.1) |  |
| Prefer not to disclose | 1 (0.1) | 1 (0.1) |  |
| **Education** |  |  |  |
| Intermediate or below | 119 (24.9) | 188 (20.6) | 0.069 |
| Bachelor's or above | 358 (74.9) | 721 (79.0) |  |
| Prefer not to disclose | 1 (0.2) | 4 (0.4) |  |
| **Household incomes** |  |  |  |
| PKR 20,000 or below | 33 (8.4) | 98 (12.0) | 0.877 |
| PKR 20,001 – PKR 40,000 | 53 (13.5) | 133 (16.2) |  |
| PKR 40,001 – PKR 60,000 | 40 (10.2) | 99 (12.1) |  |
| PKR 60,001 – PKR 80,000 | 28 (7.1) | 74 (9.0) |  |
| PKR 80,001 – PKR 100,000 | 28 (7.1) | 58 (7.1) |  |
| PKR 100,001 – PKR 120,000 | 33 (8.4) | 85 (10.4) |  |
| > PKR 120,000 | 63 (16.0) | 135 (16.5) |  |
| Prefer not to disclose | 115 (29.3) | 137 (16.7) |  |
| **Permanent residence** |  |  |  |
| Karachi | 329 (68.8) | 364 (39.9) | **<0.001** |
| Lahore | 36 (7.5) | 91 (10.0) |  |
| Islamabad | 25 (5.2) | 56 (6.1) |  |
| Peshawar | 8 (1.7) | 45 (4.9) |  |
| Quetta | 2 (0.4) | 13 (1.4) |  |
| Hyderabad | 18 (3.8) | 34 (3.7) |  |
| Others* | 60 (12.6) | 310 (34.0) |  |

\*\*Percentages may not total 100 due to rounding
‡Pearson Chi-Square test

Around three-fourths of respondents perceived that they (strongly agree/agree: 71% women vs. 73% men) and their family (strongly agree/agree: 74% women vs. 73% men) might be infected with COVID-19 if no preventive measures were taken. However, significantly more women than men considered symptoms of COVID-19 (if infected) as severe (very severe/severe: 46% women and 39% men, p-value: 0.045). On probing regarding, 59% women vs. 73% men perceived themselves likely to survive an infection (Table 2).

**Table 2.**
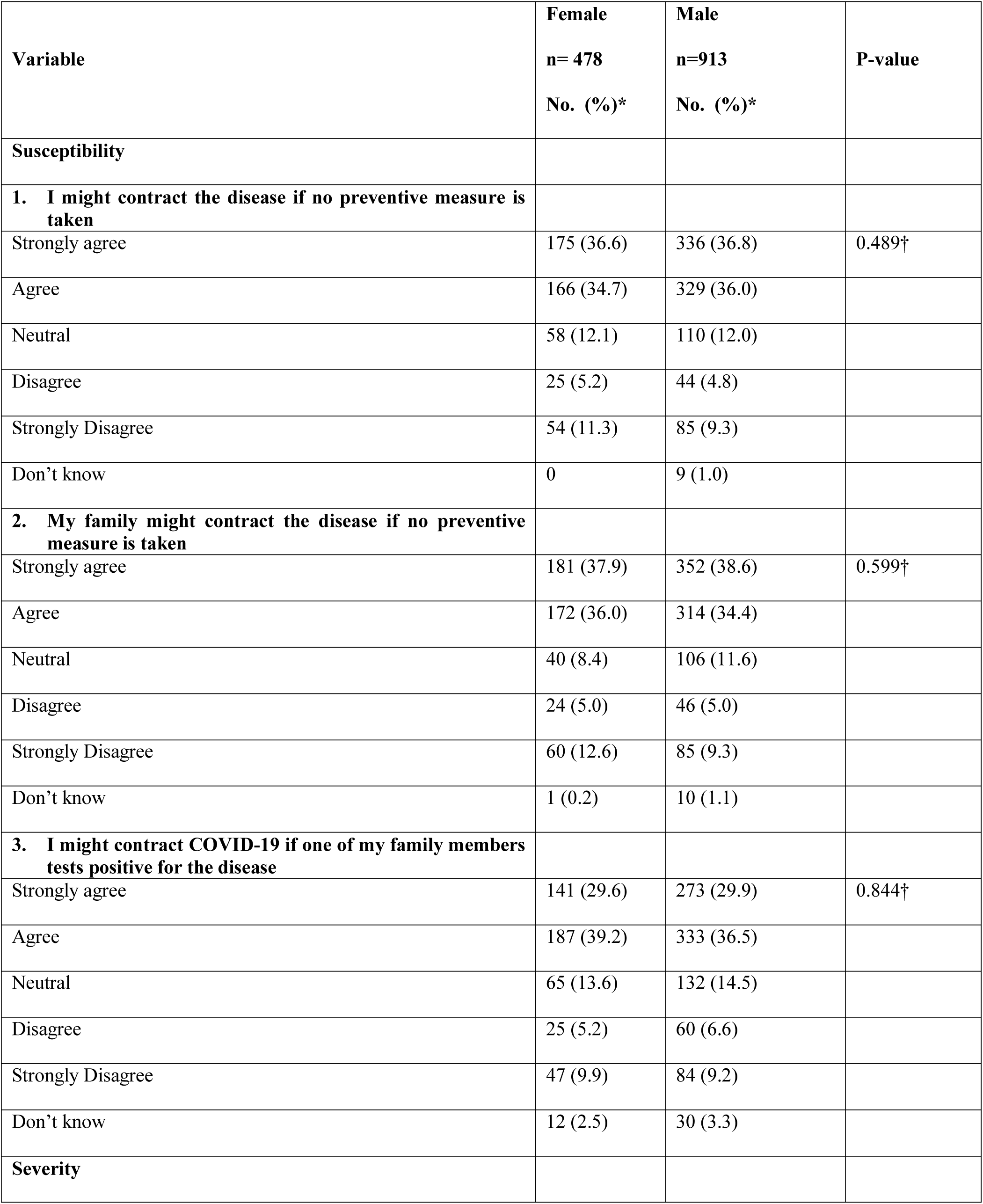
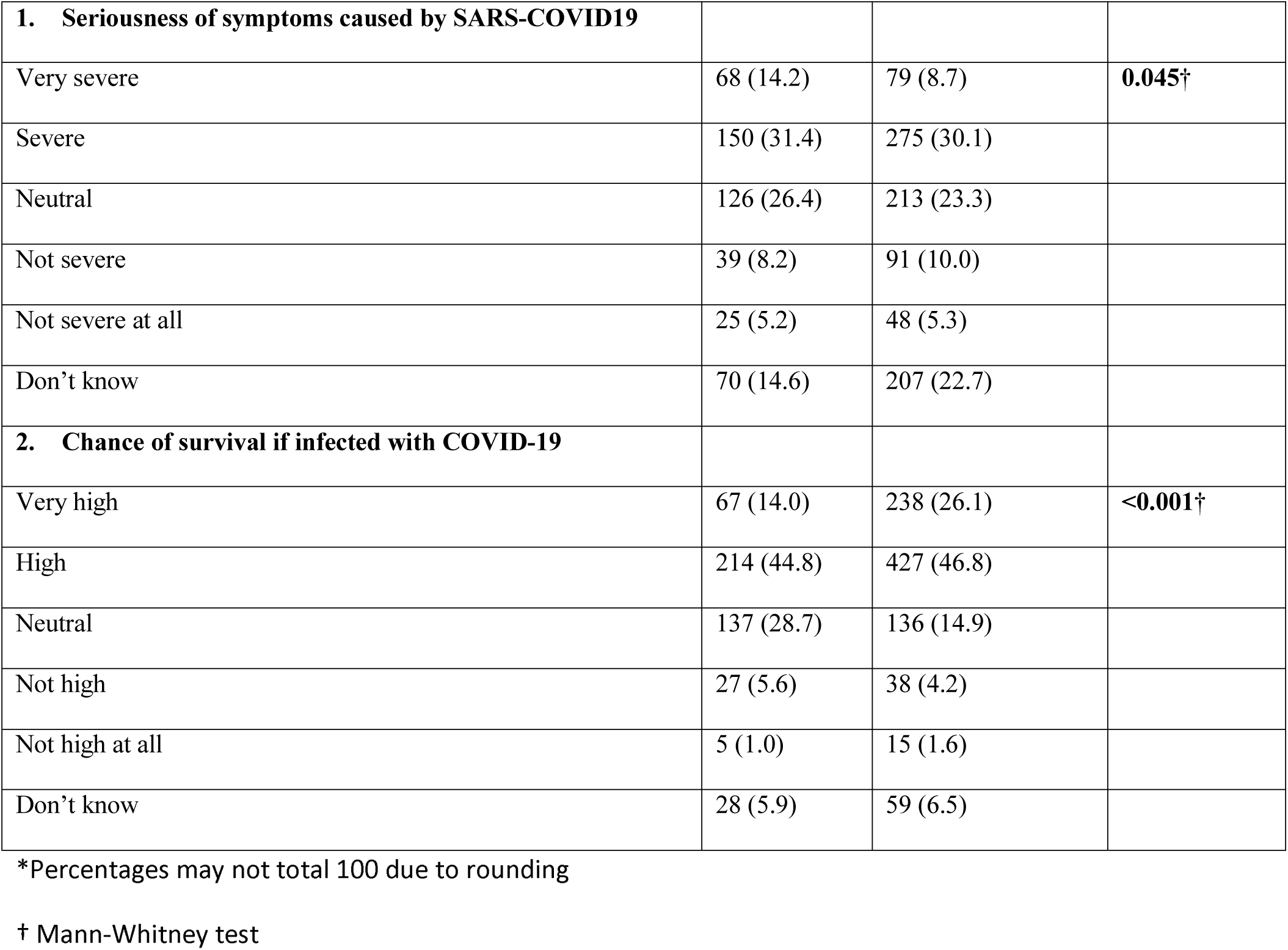
Perceived Severity and Susceptibility for COVID-19 in women and men.

Women were also reported to have a higher anxiety score (Mean ± SD: 6.80 ± 3.61 in women, and 5.93 ± 3.58 in men, p-value: <0.001).. Furthermore, the depression score was high among women (Mean ± SD: 8.39 ± 3.93 in women 8.01 ± 3.69 in men, p-value: <0.079). More women were found to be depressed compared to men, with 58% of women and 54% of men scoring above the depression cut-off point(≥ 8).Around three-fifths of the respondents (strongly agree/agree: 58% women and 61% men) mentioned that COVID-19 had affected their jobs.

About three-fourths of the respondents (strongly agree/agree: 73% women and 74% men) also expressed concerns that the current pandemic is impacting their personal life. About two-fourths of the respondents believed that their sleeping pattern (strongly agree/agree: 40% women and 39% men) and eating habits (strongly agree/agree: 36% both women and men) have been disturbed due to COVID-19. Significantly more men compared to women mentioned that they might start/increase cigarette consumption (strongly agree/agree: 6% women vs. 11% men, p-value: <0.001), and might resort to the use of recreational drugs such as marijuana, crystallised methamphetamines, cocaine or opium products etc. (strongly agree/agree: 4% women vs. 6% men) (Table 3).

**Table 3.**
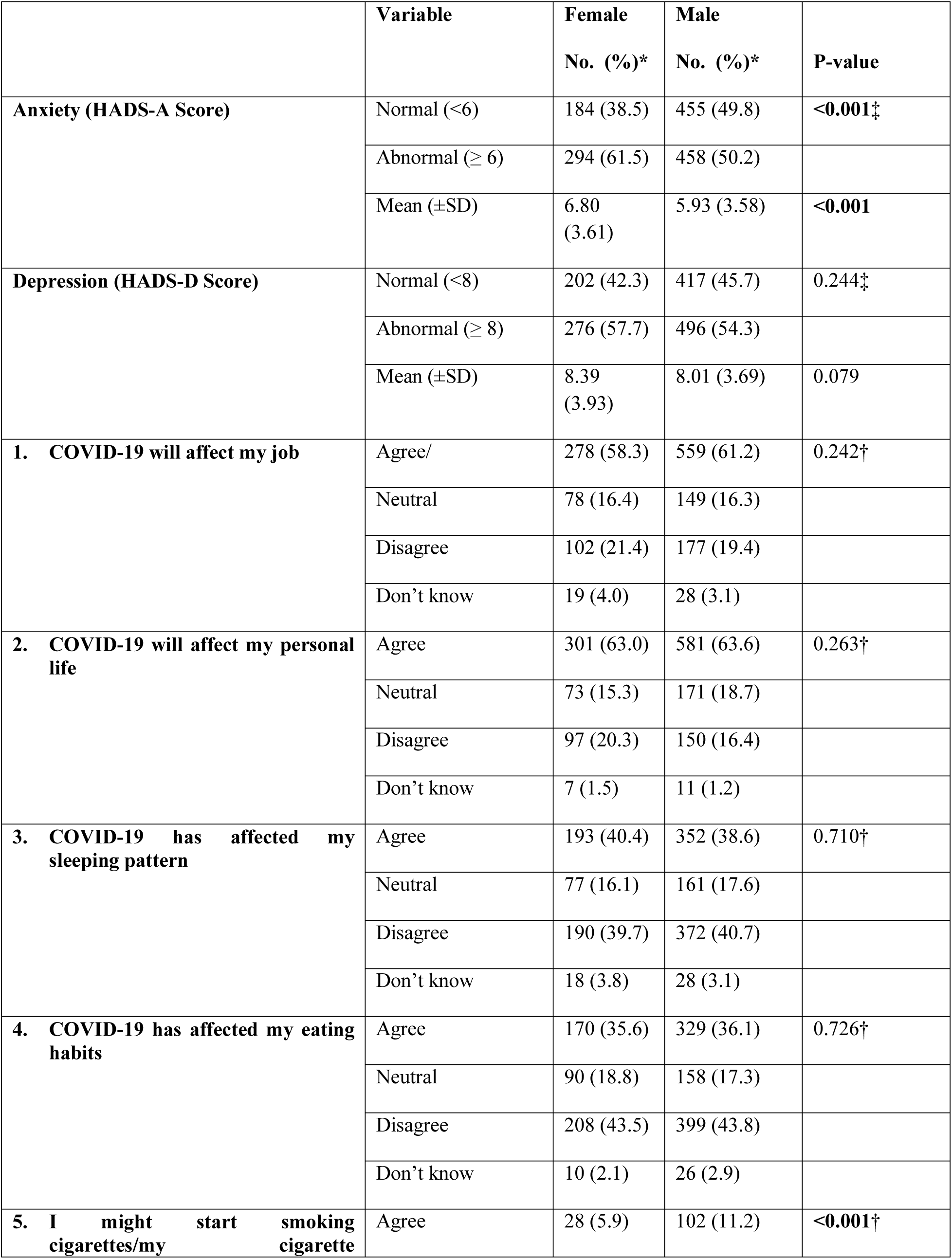
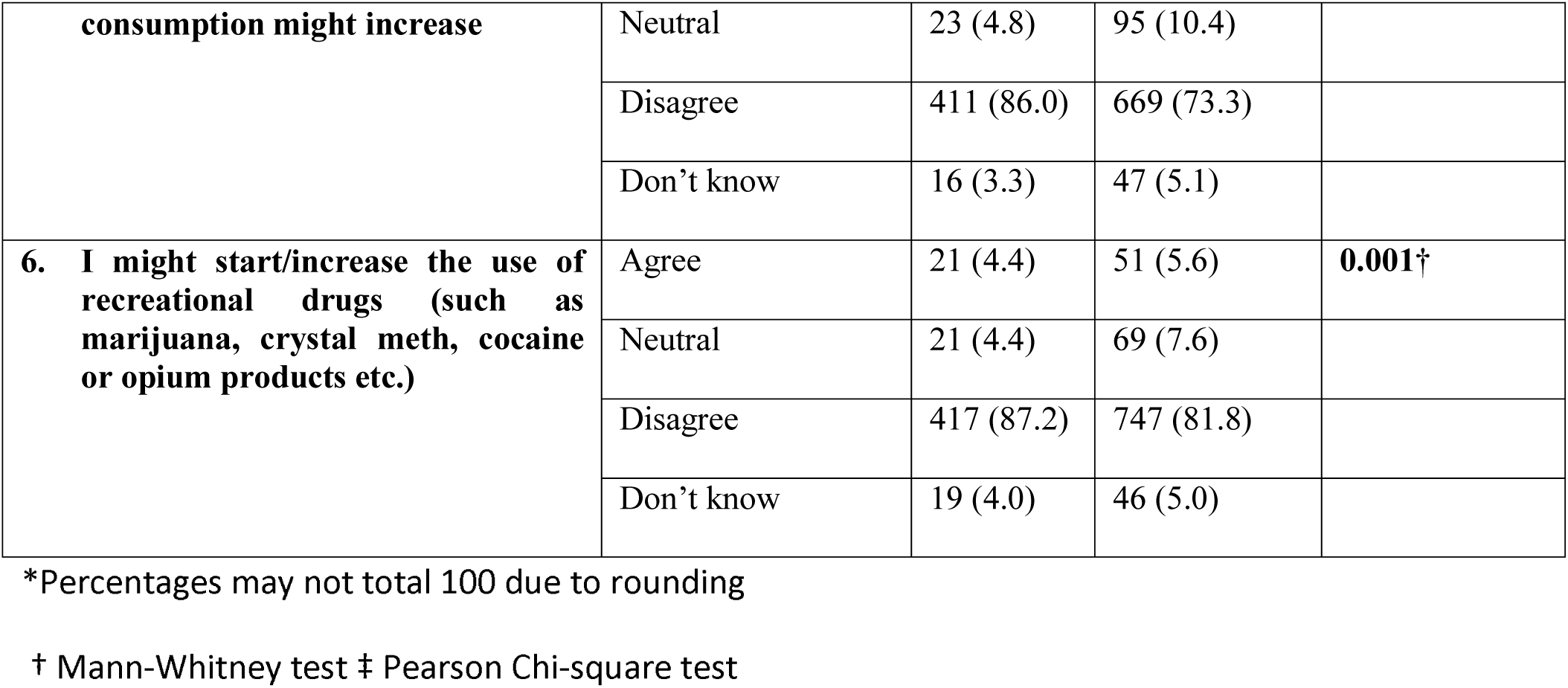
Psychological Impact ofCOVID-19 among men and women.

Significant differences were identified between women and men in adopting several precautionary measures such as washing their hands with soap/sanitizer frequently (100% women vs. 98% men, p-value: 0.012), wearing masks (93% women vs. 92% men, p-value: 0.025), covering nose and mouth while sneezing or coughing (98% women vs. 95% men, p-value: 0.043), avoiding contacting people who have fever or respiratory symptoms (95% women and 91% men), avoiding going out (87% women vs. 71% men, p-value: <0.001), avoiding crowded areas (96% women and 92% men, p-value: 0.003), refraining from going to hospital or clinic (78% women and 71% men, p-value: <0.001), avoiding to go to work (72% women and 57% men, p-value: <0.001), avoiding social events (97% women and 93% men, p-value: 0.046), and avoiding domestic travel (93% women and 86% men, p-value: <0.001). (Table 4.)

**Table 4.** Adoption of Precautionary measures.

| Variable | Female<br>No. (%) | Male<br>No. (%) | P-value |
| --- | --- | --- | --- |
| **Adoption of Precautionary Measures (frequency of yes response) \*** |  |  |  |
| 1. Wear face masks | 444 (92.9) | 834 (91.3) | **0.025**‡ |
| 2. Wash hands frequently (With soap or hand sanitizer) | 476 (99.6) | 891 (97.6) | **0.012**‡ |
| 3. Disinfecting floors and tables at home (with phenyl products) | 391 (81.8) | 646 (70.8) | **<0.001**‡ |
| 4. Cover nose and mouth when sneezing or coughing | 468 (97.9) | 870 (95.3) | **0.043**‡ |
| 5. Avoid contacting people who have fever or respiratory symptoms | 452 (94.6) | 832 (91.1) | **0.040**‡ |
| 6. Avoid contacting people who have been traveling abroad within one month | 441 (92.3) | 800 (87.6) | 0.058‡ |
| 7. Avoid going out | 417 (87.2) | 646 (70.8) | **<0.001**‡ |
| 8. Avoid crowded areas | 457 (95.6) | 840 (92.0) | **0.003**‡ |
| 9. Avoid going to meat shops/market | 391 (81.8) | 580 (63.5) | **<0.001**‡ |
| 10. Avoid going to hospital or clinic | 373 (78.0) | 648 (71.0) | **<0.001**‡ |
| 11. Avoid going to work | 346 (72.4) | 516 (56.5) | **<0.001**‡ |

\*Multiple answers † Mann-Whitney test ‡ Pearson Chi-square test

Information about COVID-19 provided by the doctor was considered reliable by significantly more women compared to men (very reliable/reliable: 91% women and 88% men, p-value: 0.041). Most of the respondents (very reliable/reliable: 81% women and 82% men) thought that the information provided through official websites such as those run by the government is reliable. Significantly more men than the women believed that the radio (very reliable/reliable: 46% women vs. 55% men, p-value: 0.014), and family or friends (very reliable/reliable: 46% women vs. 55% men, p-value: 0.003) are reliable sources for gaining information about COVID-19. Furthermore, television (very reliable/reliable: 57% women vs. 61% men), newspaper (very reliable/reliable: 56% women vs. 58% men), magazine (very reliable/reliable: 39% women and 44% men), social media such as Facebook, WhatsApp, Instagram (very reliable/ reliable: 28% women vs. 32% men) and unofficial websites (very reliable/reliable: 22% women and 31% men) were considered as reliable information sources by more men than women. (Table. 5)

**Table 5.** Perceived reliability of information sources in Pakistan by Gender.

|  | Female | Male |  |
| --- | --- | --- | --- |
| Variable | n= 478 | n=913 | P-value† |
|  | No. (%)* | No. (%)* |  |
| **Reliability of information sources** |  |  |  |
| **a. Newspaper** |  |  |  |
| Reliable/Very reliable | 248 (56.0) | 512 (58.2) | 0.843 |
| Neutral | 150 (33.9) | 256 (29.2) |  |
| Unreliable/Very unreliable | 45 (10.1) | 111 (12.6) |  |
| **b. Magazine** |  |  |  |
| Reliable/Very reliable | 173 (38.5) | 389 (44.0) | 0.104 |
| Neutral | 186 (41.4) | 329 (37.2) |  |
| Unreliable/Very unreliable | 90 (20.0) | 167 (18.9) |  |
| **c. Radio** |  |  |  |
| Reliable/Very reliable | 207 (46.0) | 486 (54.9) | **0.014** |
| Neutral | 185 (41.1) | 284 (32.1) |  |
| Unreliable/Very unreliable | 58 (12.9) | 115 (13.0) |  |
| **d. Television** |  |  |  |
| Reliable/Very reliable | 258 (57.3) | 541 (61.1) | 0.401 |
| Neutral | 117 (26.0) | 206 (23.3) |  |
| Unreliable/Very unreliable | 75 (16.7) | 138 (15.6) |  |
| **e. Official websites, like the Government** |  |  |  |
| Reliable/Very reliable | 367 (81.4) | 727 (82.2) | 0.507 |
| Neutral | 61 (13.5) | 93 (10.5) |  |
| Unreliable/Very unreliable | 23 (5.1) | 64 (7.2) |  |
| **f. Unofficial websites** |  |  |  |
| Reliable/Very reliable | 99 (22.0) | 271 (30.7) | **0.013** |
| Neutral | 138 (30.7) | 256 (29.0) |  |
| Unreliable/Very unreliable | 212 (47.3) | 357 (40.3) |  |
| **g. Social media platforms (WhatsApp, Facebook, Instagram)** |  |  |  |
| Reliable/Very reliable | 131 (29.2) | 286 (32.3) | 0.459 |
| Neutral | 130 (29.0) | 236 (26.6) |  |
| Unreliable/Very unreliable | 188 (41.8) | 364 (41.1) |  |
| **h. Your doctor** |  |  |  |
| Reliable/Very reliable | 410 (91.1) | 775 (87.6) | **0.041** |
| Neutral | 33 (7.3) | 96 (10.8) |  |
| Unreliable/Very unreliable | 7 (1.6) | 14 (1.6) |  |
| **i. Your family or friends** |  |  |  |
| Reliable/Very reliable | 206 (46.0) | 471 (53.3) | **0.003** |
| Neutral | 155 (34.6) | 288 (32.6) |  |
| Unreliable/Very unreliable | 87 (19.4) | 125 (14.1) |  |

\*Percentages may not total 100 due to rounding
† Mann-Whitney test

Table 6 demonstrates the predictors of anxiety. Gender, age, and household income had a significant positive association with anxiety. Women were nearly two times more likely to be anxious than men (aOR: 1.70, 95% CI: 1.26-2.28). Moreover, respondents of a younger age (25-34 years) (aOR 2.30, 95% CI: 1.26-4.18) were nearly two times more likely to have anxiety than respondents above 55 years of age. Respondents with a household income between PKR 60,000 and PKR 120,000 were more likely to have anxiety than respondents with a household income of >PKR 120,000 (aOR: 1.84; 95% CI: 1.27-2.67).

**Table 6.** Predictors of Anxiety in Pakistan.

| Characteristics | cOR (95% CI) | P-value | aOR* (95% CI) | P-value |
| --- | --- | --- | --- | --- |
| **Gender** |  |  |  |  |
| Female | 1.59 (1.27-1.99) | **<0.001** | 1.69 (1.26-2.27) | **<0.001** |
| Male | Reference |  | Reference |  |
| **Age** |  |  |  |  |
| 18-24years | 2.36 (1.65-3.38) | **<0.001** | 2.68 (1.70-4.23) | **<0.001** |
| 25-34 years | 2.57 (1.80-3.68) | **<0.001** | 2.77 (1.82-4.22) | **<0.001** |
| 35-44 years | 2.42 (1.64-3.56) | **<0.001** | 2.53 (1.61-3.99) | **<0.001** |
| 45 or above | Reference |  | Reference |  |
| **Household incomes** |  |  |  |  |
| ≤ PKR 60,000 | 1.69 (1.20-2.36) | **0.002** | 1.49 (1.05-2.11) | **0.027** |
| PKR 60,001 – PKR 120,000 | 1.85 (1.29-2.65) | **0.001** | 1.84 (1.27-2.67) | **0.001** |
| > PKR 120,000 | Reference |  | Reference |  |

\*Adjusted for gender, age, education, household income

Table 7 shows the predictors of depression. Only household income was found have a significantly positive association with depression in multivariate analysis. Respondents having a household income of PKR 60,000 – PKR 120,000 were more likely to have anxiety in comparison with respondents who had a household income of >PKR 120,000 (aOR: 1.99; 95% CI: 1.38-2.87).

**Table 7.** Predictors of Depression in Pakistan.

| Characteristics | cOR (95% CI) | P-value | aOR (95% CI) | P-value |
| --- | --- | --- | --- | --- |
| **Gender** |  |  |  |  |
| Female | 1.15 (0.92-1.44) | 0.224 | 1.28 (0.96-1.71) | 0.087 |
| Male | Reference |  | Reference |  |
| **Age** |  |  |  |  |
| 18-24years | 1.48 (1.04-2.09) | **0.027** | 1.26 (0.81-1.95) | 0.305 |
| 25-34 years | 1.34 (0.95-1.89) | 0.093 | 1.32 (0.88-1.97) | 0.183 |
| 35-44 years | 1.42 (0.97-2.07) | 0.069 | 1.44 (0.93-2.24) | 0.103 |
| 45 or above | Reference |  | Reference |  |
| **Household incomes** |  |  |  |  |
| ≤ PKR 60,000 | 1.57 (1.12-2.19) | **0.009** | 1.53 (1.09-2.17) | **0.015** |
| PKR 60,001 – PKR 120,000 | 1.99 (1.39-2.85) | **<0.001** | 1.99 (1.38-2.87) | **<0.001** |
| > PKR 120,000 | Reference |  | Reference |  |

\*Adjusted for gender, age, education, household income,

## Discussion

This study assessed how gender roles in Pakistan can impact anxiety levels and behavioural responses among men and women during the COVID-19 pandemic. Both men and women were found to be mildly anxious due to the COVID-19 pandemic. However, compared to men, more women perceived the disease to be fatal, and were more likely to engage in preventive behavior. These results highlight a greater need to develop gender-responsive policies in the fight to contain COVID-19.

Overall, fewer women than men responded to the questionnaire. This may be due to a prominent male dominated access to internet facilities in Pakistan. Indeed, several reports on internet penetration found that, at least three-fourths of internet and social users in the country are male (25). In Pakistan, cellular devices remain the most frequent means of accessing internet facilities, and there is a gender gap of 38%in mobile phone ownership (26).

There were also significant differences in respondents’ city of permanent residence. Three-fifths of all the female participants, and two-fifths of the males were from Karachi. This was expected, as the survey tool was disseminated over a Karachi-based university hospital’s Facebook page. The remaining respondents were from Punjab and Sindh. There are significant provincial disparities in access to internet facilities. Islamabad Capital Territory has the highest internet penetration, followed by Punjab and Sindh (27). The fewest respondents were from Balochistan, further reflecting the province’s poor internet accessibility. One-third of the male respondents were from smaller cities and towns throughout the country, compared to one-tenth of women. Differences in the nature of Pakistani men in smaller towns/cities may impact mobile phone ownership and social media usage, and explain why fewer women were from these towns and cities (28).

Although men and women considered themselves equally susceptible to a COVID-19 infection, women were more likely to perceive the disease to be fatal. This is a misperception, as gender-disaggregated data (until 24 June) on COVID-19 in Pakistan shows that three-quarters of diagnosed cases and deaths were among males, compared to a quarter among females (29). Gender-specific patterns of smoking are implicated as a significant contributor to disease severity among men (30). Indeed, this study also suggested that men were twice as likely as compared to women in reporting that either they might start smoking cigarettes and using recreational drugs or their usage might increase. These gender disparities in use of tobacco and narcotics are frequently seen in Pakistan (31–33). Furthermore, men are more likely to suffer from non-communicable diseases (34). Research suggests that excess mortality during the pandemic is higher than if these surplus deaths were caused by COVID-19 alone, particularly among cancer or heart disease patients and are attributable to delays in seeking or obtaining lifesaving care (35,36). Furthermore, women, particularly working mothers, tend to spend more time than men focused on medical issues related to their family’s healthcare, as well as their own (37). This could explain why women are more likely to believe that the disease symptoms are severe, with a low likelihood of survival.

Despite at being lower risk, women were more likely to conform to preventive measures. This included practicing social distancing measures, such as avoiding going to meat shops/market, going out, and going to work. One digital ethnographic study suggests that the majority of the population in Pakistan was in favor of continuing prayers in mosques while 1 in 4 men reported to have attended Friday prayers (38). Other studies also demonstrate men’s priorities during the pandemic. A comparison of COVID-19-related content shared on Twitter by men and women based in the U.S. found that women were more likely to tweet about family, social distancing and healthcare whereas men were more likely to tweet about sports cancellations and politics (39).

While this finding might imply that men are considerably less interested in social distancing practices, only one-fifth of the Pakistani women contribute to the labor force and wherein men constitute a majority of waged and salaried workers in cities and women contribute to over 70% of share of work in agriculture and informal sectors (40).These differences in employment could explain why men are less likely to conform to social distancing practices than women. In order to improve the labor forces’ capacity to work from home, initiatives should be taken to improve telecommunication facilities (including improving internet service provision).

Differences were also seen in practicing hygiene measures, such as disinfecting floors and tables at home (with phenyl products). School closure, lockdown and work-from-home orders have resulted in women carrying a double shift of home-schooling and working responsibilities. This increases the proportion of paid and unpaid labour in women’s work (41). Working mothers spend more hours engaged in household work and child care than their husbands (42). One study conducted in the United Kingdom during the lockdown estimates that, on average, mothers spend 11 more hours weekly on childcare than fathers. Single parents have less time to spend on childcare than partnered mothers, as they are single-handedly forced to bear the brunt of the shifts in the job market (43). This additional housework could result in women permanently exiting from the labor market. These developments are concerning, and emphasize the urgent need to develop labour policies which protect women in the workforce.

Women were more likely to report that they avoid going to hospitals. Furthermore, they were also more likely to perceive that doctors were a reliable source of information. It may be too soon to estimate the impact of COVID-19 on maternal and child health services, but one study estimates that a modest decline of 10% in coverage of pregnancy-related and newborn health care in lower-middle income countries could result in an additional, 1.7 million women who give birth, and 2.6million newborns who need urgent medical care (44). Studies conducted during the 2013–2016 Ebola outbreak in Western Africa show how sexual and reproductive health was adversely impacted by strains on health care systems, which often resulted in interruptions of care, and redirected resources (45–49). Similar reduction in access can be seen during the current pandemic. Clinics operated by Marie Stopes International, which is the largest private provider of family planning services in India, had to halt operations due to the country-wide lockdowns (50). Similarly, Marie Stopes International reports that its activities have been reduced by up to 40% in Pakistan due to the pandemic (51). Furthermore, some studies noted how diversion of staff and funding from maternal, neonatal and child health programmes to the front-line of the COVID-19 response has also decreased the quality of services available to women (52–54)

A larger number of men than women considered radio to be a reliable source of information. Pakistan has a considerable audience of radio programming. One study found that radio has the largest listeners in Sindh (60%), followed by Balochistan (53%), KPK (52%) and Punjab (19%), particularly in rural areas and small towns (55). Therefore, this gendered difference in trusting radio could be explained by the significantly larger proportion of men who came from smaller cities and towns. Furthermore, men in this study were more likely to trust friends and family than women. Pakistanis are considered to be a collectivistic society, with an emphasis on men engaging in commitments to the members of their ‘group’, friends and family (56). Similarly, men were more likely to trust social media sources than women. The lockdown enforced due to COVID pandemic has resulted in online activity substituting social activity between families, and may be considered as representative of a given person’s public interactions (38,39). While one study found that the public was often skeptical of official figures on COVID-19, but most polling suggests that Pakistanis in general are confident in the government’s management of the crisis (4,5). This is in contrast with Syria, where experiences of war, and propaganda campaigns by the state and its opponents, meant that Syrians were very distrustful of official news sources (38). Facebook users were much more likely to share official news sources. Therefore, radio content, as well as shareable social media content might be an appropriate avenue to provide targeted health information to men and improve their risk perceptions and subsequent indulgence in precautionary measures

In this study, women depicted higher levels of anxiety and depression in comparison with men, which suggests that they hold a greater psychiatric burden of the COVID-19 pandemic. One study conducted in India found that 33% of the respondents had experienced either depression or anxiety as a result of COVID-19 (57). Another study in China established that 54% of respondents suffered some psychological impact from the outbreak (58). In both studies, women were found to have suffered a greater psychological impact due to the pandemic as compared to men. Similarly, our findings corroborate with data from Turkey, where women had significantly higher scores of depression and anxiety (14). Research conducted in China also reports that women may be three times more anxious than their male counterparts because of COVID-19 (11,54,58). These results align with previous studies which show that women have a higher vulnerability for developing anxiety disorders (59)

There are many possible sources of this concern. Apart from their professional role, women serve as primary caregivers within their family (57). Women’s greater sensitivity towards familial roles and responsibilities was also reflected in a research which noted that pregnant women had heightened stress levels regarding the health status of their older relatives, their children and then their unborn baby during COVID-19 (60). One study found that family income stability and social support networks were protective factors against anxiety (61). Given the difficulties in forming stable social support networks during pandemic-induced lockdowns, as well as women’s lower likelihood of seeking medical care, this psychological support could be provided via telemedicine or other online platforms which can connect them to qualified psychiatrists. The timely and effective provision of such psychological support is also imperative for literate women that are already suffering from mental illness and feel that their symptoms have aggravated due to the ongoing pandemic.

## Conclusion

This study assessed the gender differences in risk perceptions (susceptibility and severity of the disease), preventive behaviour (social distancing, enhanced hygiene measures), and anxiety. The results highlight the need for gender-responsive policies in mitigating the health and economic impact of the COVID-19 pandemic. The global economy has grinded to a stop, and respondents face severe economic uncertainty. Differences in type-of-work, based on gender, may result in men being unable to maintain social distancing. Furthermore, it results in women being burdened with increasing house work, which can impact their ability to engage in professional work. This indicates the urgent need to develop labour laws to protect the workforce, particularly women. Furthermore, the results indicate potential avenues of disseminating gender-specific health communication. Involving doctors in healthcare communication targeting women, focusing on their need to avoid skipping hospital appointments, might prove effective. Research is required to assess strategies of reducing the frequency of in-person MNCH appointments, and the potential of telemedicine in all women to remain in contact with the health system, As men are more likely to trust what they read on social media, especially if it is shared by friends or family, social media campaigns and radio programming may be effective in disseminating information and the latter could be an effective tool to reach towns and cities. This health communication should include messages about men’s higher risk of dying, due to COVID-19, a lack of NCD management, and smoking cessation. Most importantly, based on the discussion, policy measures must be taken to ensure the continued provision of quality healthcare to women. This must include provisions to mitigate the growing anxiety among women, and compensate for the loss of social support networks during pandemic times.

Differences in type-of-work, based on gender, may result in men being unable to maintain social distancing. Furthermore, it results in women being burdened with increasing house work, which can impact their ability to engage in professional work.

## Supporting information

STROBE checklist for cross-sectional studies

## Data Availability

Data will be made available from the corresponding author on reasonable request.

## Author Contributions

FR conceived the study, guided data collection and reviewed all drafts of the manuscript. HAK and ARK adapted the questionnaires and wrote the manuscript. SP analyzed and narrated the data. HAK and ARK edited and revised multiple drafts of the manuscript. ARK also assisted in adapting the questionnaires and posting it on social media channels of AKU. All authors reviewed and endorsed the final submission.

## Acknowledgments

The authors would like to thank all the respondents of the survey from across Pakistan

